# Establishment of Contextually Appropriate Cut Offs for Orthopoxvirus Serologic Assays in an Mpox-Endemic Setting

**DOI:** 10.64898/2026.04.10.26350607

**Authors:** Christina Frederick, Sydney Merritt, Megan Halbrook, Patrick Mukadi, Yvon Anta, Jean Paul Kompany, Merly Tambu, Jean Claude Makangara-Cigolo, Emmanuel Hasivirwe Vakaniaki, Michel Kenye, Lygie Lunyanga, Cris Kacita, Thierry Kalonji, Christophe Kinanga, Sylvie Linsuke, Lisa E. Hensley, Isaac I. Bogoch, Souradet Y. Shaw, Nicole A. Hoff, Placide Mbala-Kingebeni, Anne W. Rimoin, Jason Kindrachuk

## Abstract

Mpox virus (MPXV) gained increased attention following the declaration of two Public Health Emergencies of International Concern (PHEICs) in 2022 and 2024. The rapid spread of MPXV and the increase in human-to-human transmission highlighted the need for improved diagnostic tools for characterizing infection patterns and transmission dynamics. While PCR is effective for detecting active infections, serological approaches can help identify previous or asymptomatic infections and support retrospective surveillance. However, many serological assays developed during recent outbreaks have not been evaluated in endemic settings such as the Democratic Republic of the Congo (DRC). This study aims to define antigen-specific serological cutoff values to differentiate MPXV-seroreactive individuals from those with other orthopoxvirus (OPXV) exposure or different vaccination histories, specifically for use in the DRC. Here, we analyzed 134 individuals, divided into six distinct cohorts with different exposures. Serum samples were tested using Mesoscale Discovery (MSD) to screen for five MPXV and vaccinia virus (VACV) orthologous antigens: A29L/A27L, A35R/A33R, B6R/B5R, E8L/D8L, and M1R/L1R. Receiver operating characteristic (ROC) analysis identified the best-performing antigens and established seroreactivity cutoff values. A binary composite rule was also evaluated to improve the classification of these results. We identified three MPXV antigens, E8L (cut-off=12.33 AU/mL), A35R (cut-off=5.22 AU/mL), and B6R (cut-off=9.77 AU/mL), that showed the strongest discriminatory performance in the dataset. Collectively, these three antigens form a significant panel that demonstrated clear separation between our mpox survivor cohort and other OPXV-exposed individuals.

**Importance:** Establishing antigen-specific serological cutoff values for this assay using unique samples from endemic regions such as the DRC may improve future epidemiological and disease transmission surveillance efforts and contribute to broader efforts to ensure regionally appropriate cutoffs for serological assays.

## BACKGROUND

Mpox virus, also known as monkeypox virus (MPXV), is a zoonotic pathogen belonging to the *Poxviridae* family and the *Orthopoxvirus* (OPXV) genus. MPXV has been circulating endemically in Central and West African countries for several decades, most notably the Democratic Republic of the Congo (DRC) [1–5]. MPXV is classified into two major clades. Clade I, which is endemic in forested regions of Central Africa, is subdivided into Clade Ia, characterized primarily by zoonotic spillover events, and Clade Ib, characterized by sustained human-to-human transmission. Clade II MPXV, endemic in forested regions of West Africa, is similarly subdivided into Clade IIa and Clade IIb, with Clade IIb typified by sustained human-to-human transmission chains [1, 4]. Clade I and Clade II MPXV were responsible for declarations of Public Health Emergencies of International Concern (PHEIC) for mpox by the World Health Organization (WHO), in July 2022 and August 2024, respectively [4, 6, 7]. The 2022 PHEIC was driven by the rapid international spread of MPXV Clade IIb, with infections concentrated among dense sexual networks [4–6]. The 2024 PHEIC followed the emergence of the newly identified Clade Ib lineage in South Kivu, DRC, and rapid expansion across the DRC and into neighboring non-endemic countries, including Uganda, Kenya, and Burundi. The sustained human-to-human transmission dynamics of both Clade I and Clade II MPXV are also characterized by an increased rate of APOBEC3-driven mutations [4, 8]. In late August 2025, the WHO announced that mpox was no longer classified as a PHEIC; however, the virus continues to be monitored and remains an important global health concern [9].

MPXV is a double-stranded DNA (dsDNA) virus with a genome of approximately 197 kilobase pairs (kb) and a virion size of around 200–250 nm [1, 3]. The MPXV genome is linear and consists of two key components: the inverted terminal repeats (ITRs), which are approximately 10 kb each and located at both ends of the genome. ITRs are important in the MPXV genome as they direct immunomodulatory host-range factors and serve as critical identifiers for lineage characterization. In fact, both Clades I and II significantly differ in the ITR region [4, 10, 11]. The central region is conserved across both clades and contains housekeeping genes essential for viral replication and core functions. Importantly, MPXV proteins are released into the host cell with the help of Intracellular Mature Virions (IMV) and Extracellular Enveloped Virions (EEV). These components carry several viral surface proteins that facilitate attachment, membrane fusion, and entry, and include viral proteins A35R, B6R, E8L, M1R and A29L [4, 10, 11].

Before the first documentation of human mpox, smallpox, caused by the closely related variola virus, had devastating impacts on the global population, resulting in approximately 500 million deaths during the 20th century [12]. However, widespread vaccination programs using vaccinia virus-derived vaccines resulted in the eradication of smallpox by 1980 [12]. Dryvax, a first-generation smallpox vaccine, was used during the smallpox eradication program; however, it was discontinued in the early 2000s following the licensure of ACAM2000, which had reduced contraindications. ACAM2000 was administered for rapid immunization or high-risk work settings (e.g., during field deployments, military settings) [13–16]. LC16m8 is a live-attenuated vaccinia-based smallpox vaccine developed in Japan and licensed in the 1970s, historically used during the final stages of the smallpox eradication campaign and more recently repurposed for mpox prevention [17]. More recently, a third-generation vaccine, modified vaccinia Bavarian Nordic (MVA-BN; JYNNEOS; Imvamune), has been developed [5, 13, 14]. Investigations of MVA-BN deployment during the 2022 global mpox outbreak have reported efficacy against mpox ranging from 35-75% for a single vaccine dose and 66-85% with the two-dose regimen [5, 13, 14]. Characterization of the humoral and cellular responses to MVA-BN, as well as assessment of vaccine efficacy across Clades I and II MPXV, is ongoing [18].

Confirmation of active MPXV infections relies on PCR-based diagnostic testing [19, 20]. However, the true burden of MPXV is likely much higher than reported confirmed infections due to diagnostic resource limitations, healthcare access, altered clinical presentations, and stigma [16, 19, 20]. Serological assessments can provide essential information for understanding the true burden of mpox by identifying chains of asymptomatic or unreported exposures with a population-based approach [19]. Studies have shown that VACV antibodies can persist for up to 80 years in vaccinated individuals [21]. There is also evidence that MPXV antibodies can last from months to years after infection in humans, although these may gradually decrease over time [21, 22]. Yet, as MPXV and VACV share approximately 94–98% antigenic similarity, this creates substantial challenges in distinguishing MPXV-specific antibody responses from those elicited by other orthopoxvirus exposures [13, 20]. Although several commercial ELISA-based kits have recently been developed targeting MPXV-specific antigens, such as A29L and A35R, limitations remain, particularly regarding optimal application across diverse global settings [23].

Historically, regions that are endemic for mpox have been affected by a lack of sustained surveillance programs and limited laboratory infrastructure. Additionally, the challenges of accessing remote communities, combined with the complex ongoing political and security conditions across the country, remain difficult [6, 18]. These challenges can lead to an underestimation of MPXV infections and ongoing circulation, posing a risk to neighboring populations [6, 18]. These logistical and surveillance barriers can hinder the validation of diagnostic tools in endemic settings, meaning that assays developed during outbreak responses may not always be optimized for historically endemic settings. During the 2022 mpox outbreak, several serological kits were developed to enable more specific detection across multiple viral antigens. Although these kits were only tested on a limited subset of patients, they were not evaluated in endemic mpox settings such as the DRC, where establishing serological assays specific to local populations would be critical for accurate mpox detection [23]. In fact, four commercial serology kits were developed, several of which already have pre-established seropositivity cut-off values; however, these thresholds may not directly apply to populations in endemic regions, as differences in exposure history, population risk groups, and other demographic factors must be considered to ensure accurate interpretation of serological results [23].

Multiplex platforms, including Mesoscale Discovery (MSD), have shown improved sensitivity for detecting MPXV-specific IgG, and specifically for protein E8L, which has demonstrated strong discriminatory capacity between exposed and unexposed individuals [20, 23]. Comparable patterns have been observed for the VACV ortholog, D8L, for which the MSD assay also detects robust responses among mpox-infected cohorts. In addition, B6R also appears to have good diagnostic potential, though its signal separation is less pronounced. Notably, the VACV homolog B5R exhibits a nearly identical response profile, underscoring the difficulty of resolving MPXV-versus VACV-induced immunity [20, 23]. Although individual antigens show cross-reactivity between MPXV and VACV, the use of paired homologous antigen ratios within this multiplex platform allows clearer differentiation between vaccinated, infected, and unexposed individuals. This makes the assay particularly suitable for comparative serologic analyses in populations with mixed orthopoxvirus exposure histories [24].

Given these antigen-specific differences, establishing reliable and region-appropriate serological cutoffs is important for reducing cross-reactivity, improving diagnostic specificity, and facilitating more accurate interpretation of seroprevalence patterns across vaccinated and unvaccinated populations. The need for rapid and accurate diagnostic tools remains critical even after the cessation of both PHEIC declarations, as reliable screening assays are essential for evaluating the clinical and exposure profiles of suspected mpox cases.

The objective of this study is to establish serological cutoffs for each antigen in the Orthopoxvirus MSD panel using cohorts with differential OPXV exposures from the DRC. These cutoffs may enable a more accurate identification of MPXV-seroreactive individuals in future cohort studies, and larger population-wide MPXV screening.

## MATERIALS AND METHODS

### Ethical Considerations

This study was approved by the Institutional Review Boards of the University of California, Los Angeles (IRB#23–000676), University of Manitoba (HS25837) and the Kinshasa School of Public Health, University of Kinshasa, DRC (ESP/CE/161/2024).

### Cohort Selection

To establish an analytical cut-off for this study, we used a subset of serum samples collected from June 2024 to September 2024 (n = 134) from individuals recruited for a previous study. These samples were collected from Kinshasa Province, Mbandaka city (Equateur Province), and Kinsantu city (Kongo Central Province) in the DRC (Table 1). Specifically, samples were stratified into six major groups: (1) a control group (*n* = 22), consisting of individuals with no documented history of OPXV infection or vaccination from an area that had never reported confirmed mpox cases; (2) an mpox survivor group (*n* = 21), composed of participants with laboratory or clinically confirmed prior mpox infection who had subsequently recovered and were at least four weeks post infection; (3) a smallpox survivor and smallpox vaccination group (*n* = 22), representing individuals who had survived a prior smallpox infection and as a part of the mass campaigns during the final smallpox eradication period also received a smallpox vaccine; (4) a smallpox vaccination-only group (*n* = 25), comprising individuals with known vaccination history of being vaccinated prior to 1980 with Dryvax but no record of mpox or smallpox infection; (5) a JYNNEOS vaccination group (*n* = 22), consisting of participants with verified documentation of JYNNEOS vaccine administration; and (6) a JJ Ebola vaccination with MVA boost group (*n* = 22), representing individuals who had previously received the two-dose Johnson & Johnson Ebola vaccine which included an Modified VACV (MVA)-based booster. All smallpox vaccination histories were confirmed by visual identification of scars at the time of interview; historical smallpox survivors were identified in collaboration with the DRC Ministry of Health documentation, supplemented by participants’ referrals. In addition, demographic data were recorded, and participants consented to the use of their samples and information for future research. At the time of participation, no participants reported ongoing symptoms or other health concerns.

**Table 1:** Mean antigen-specific concentrations (AU/mL) across the selected cohort groups measured by Mesoscale Discovery Assay (MSD)

|  | Participants & Sample Location | Control (n=22), Kisantu | JJ Ebola Vaccination & MVA Boost (n=22), Kinshasa | Jynneos Vaccination (n=22), Kinshasa | Mpox Survivor (n=21), Kinshasa | Smallpox Survivor & Smallpox Vaccination, (n=22) Kinshasa | Smallpox Vaccination, (n=25) Mbandaka |
| --- | --- | --- | --- | --- | --- | --- | --- |
| Antigens | VACV A27L (95% CI) | 2.63<br>(1.21-4.04) | 3.26<br>(-0.55-7.08) | 2.34<br>(1.11-3.57) | 11.12<br>(3.46-18.77) | 3.58<br>(1.48-5.67) | 1.29<br>(0.62-1.96) |
|  | VACV A33R (95% CI) | 1.31<br>(0.52-2.10) | 5.27<br>(1.72-8.82) | 7.68<br>(3.42-11.94) | 129.50<br>(83.75-175.25) | 20.86<br>(7.54-34.17) | 61.22<br>(-30.34-152.78) |
|  | VACV B5R (95% CI) | 2.06<br>(0.99-3.12) | 2.50<br>(0.74-4.26) | 6.85<br>(4.45-9.25) | 91.98<br>(63.91-120.05) | 13.10<br>(4.71-21.49) | 11.20<br>(1.56-20.84) |
|  | VACV D8L (95% CI) | 7.56<br>(-4.03-19.14) | 11.07<br>(-8.53-30.68) | 39.42<br>(22.73-56.12) | 231.30<br>(103.20-359.40) | 19.16<br>(6.74-31.58) | 2.68<br>(0.80-4.56) |
|  | VACV L1R (95% CI) | 2.20<br>(0.67-3.73) | 2.23<br>(0.66-3.79) | 2.92<br>(1.56-4.29) | 197.65<br>(-122.68-517.97) | 4.62<br>(2.67-6.56) | 3.39<br>(-0.59-7.37) |
|  | MPXV M1R (95% CI) | 2.09<br>(0.42-3.75) | 2.21<br>(0.44-3.99) | 2.45<br>(1.24-3.65) | 58.44<br>(-6.94-123.81) | 3.73<br>(2.16-5.30) | 1.55<br>(0.57-2.53) |
|  | MPXV E8L (95% CI) | 3.91<br>(0.66-7.16) | 17.01<br>(-13.64-47.66) | 22.21<br>(13.24-31.19) | 699.09<br>(460.73-937.45) | 17.56<br>(-0.74-35.86) | 2.70<br>(-0.08-5.48) |
|  | MPXV B6R (95% CI) | 1.66<br>(0.90-2.42) | 2.28<br>(0.34-4.21) | 5.97<br>(3.63-8.30) | 135.51<br>(92.61-178.40) | 14.38<br>(3.97-24.79) | 11.83<br>(0.96-22.71) |
|  | MPXV A35R (95% CI) | 2.21<br>(-0.18-4.60) | 6.40<br>(2.13-10.66) | 3.23<br>(2.17-4.28) | 504.88<br>(341.39-668.38) | 23.49<br>(7.13-39.85) | 64.41<br>(-27.34-156.16) |
|  | MPXVA29L (95% CI) | 2.77<br>(1.26-4.28) | 4.23<br>(-0.20-8.66) | 3.19<br>(1.15-5.23) | 9.87<br>(4.74-14.99) | 5.14<br>(2.03-8.25) | 1.92<br>(0.89-2.95) |

### Mesoscale Discovery Assay

Sera samples from each participant were analyzed using the MESO QuickPlex SQ120 MM instrument (Meso Scale Discovery) with the V-PLEX Orthopoxvirus IgG Serology Kit as per manufacturer instructions. The Orthopoxvirus panel includes 5 homologous pairs, notably A29L / A27L, A35R / A33R, B6R / B5R, E8L / D8L, and M1R / L1R [25]. Each well contained a unique sample, with no duplicates or serial dilutions. Raw signals were exported to Discovery Workbench 4.0 for, where antigen-specific concentrations (AU/mL) were generated. These values served as the basis for establishing antigen-specific serological cutoffs (Table 1, Figure 1).

**Figure 1:**
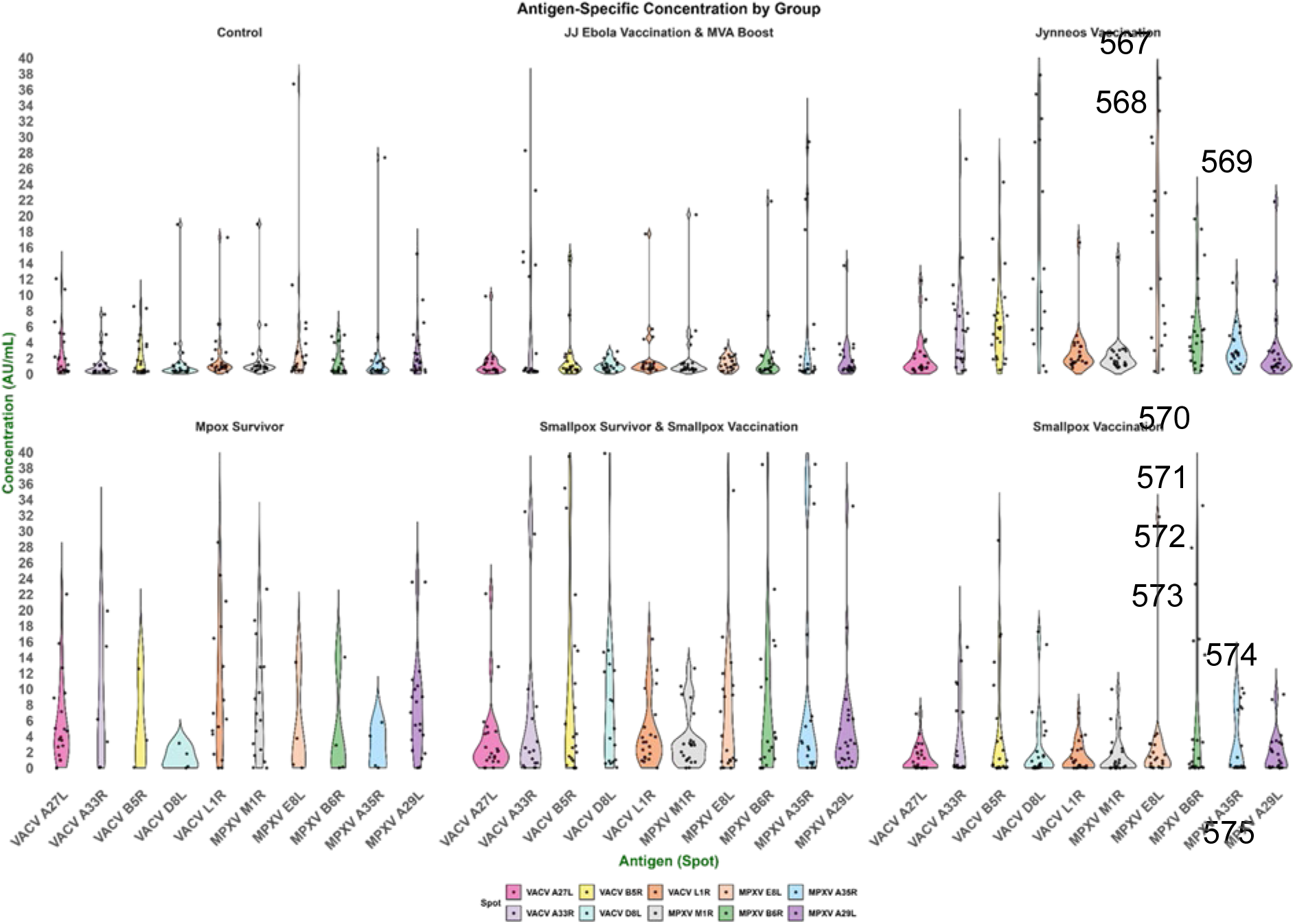
Antigen-specific concentrations (AU/mL) across the selected cohort groups measured by MSD

### Ortholog Ratios

Electrochemiluminescence (ECL) values represent the raw antigen–antibody binding signal measured in each well on the MSD platform and were used to visualize signal intensity and assess assay performance across antigens and cohorts (Figure S3, Table S2). These ECL values were then processed using MSD Discovery Workbench software to convert signals into antigen-specific concentrations (AU/mL), which were subsequently used to evaluate ROC metrics and determine cut-off values.

## RESULTS

Across all six cohorts, the mpox survivor group displays highly variable antibody reactivity compared with the other groups (Figure 1). The control and the JJ Ebola Vaccination groups show the lowest overall variability in reactivity across all antigens. Elevated variability is also observed in antibody responses to D8L/E8L and B5R/B6R among individuals in the smallpox survivor and Jynneos vaccination cohorts. In these groups, antibody responses appear more dispersed across participants. Among the control group, no antibody concentration against any antigen exceeded 8.0 AU/mL (Table 1). In contrast, the mpox survivor group showed significantly higher antibody levels, especially against VACV D8L and MPXV homolog E8L, with mean concentrations of 231.30 AU/mL and 699.09 AU/mL, respectively. Elevated responses were also observed for VACV A33R and MPXV homolog A35R, with mean concentrations of 129.50 AU/mL and 504.88 AU/mL, respectively. Except for MPXV A29L and MPXV M1R, antibody concentrations against MPXV antigens in the mpox survivor group were generally higher than those for their corresponding VACV orthologs. While Table 1 summarizes the mean antibody concentrations for each cohort, the individual-level distributions provide a clearer view of the heterogeneity of antibody responses within each group (Figure 1). This may also be influenced by different time points of infection, which could affect antibody levels in sera.

### Establishment of a ROC Curve

Optimal assay cut-off values were determined using Receiver Operating Characteristic (ROC) curve analyses, in which responses from the control group (n = 22; true negatives) were compared against those from mpox survivors (n = 21; true positives) and other vaccine-exposed individuals (n = 91). Thresholds were selected to maximize diagnostic sensitivity and specificity [26].

### ROC Curve

ROC analysis was used to evaluate the discriminative ability of MPXV and VACV-derived antigens in identifying individuals with prior mpox infection. Most antigens displayed excellent classification performance, with area under the curve (AUC) values around 0.90, reflecting strong humoral reactivity in convalescent sera. MPXV A35R (AUC = 0.91; 95% Confidence Interval (CI) = 0.79-1.00; Sensitivity (Se) = 0.86; Specificity (Sp) = 0.95; cutoff = 5.22 AU/mL) and MPXV E8L (AUC = 0.90; 95%(CI) = 0.78-1.00; Se = 0.86; Sp = 0.95; cutoff = 12.33 AU/mL) were the most discriminative antigens, followed closely by cross-reactive VACV A33R and B5R (AUC ≥ 0.90). These results indicate that A35R and E8L capture strong antibody responses to MPXV while maintaining cross-orthopox reactivity with Vaccinia homologs. Antigens such as A27L and A29L displayed moderate accuracy (AUC 0.72–0.77).

### Testing of mpox survivor cut-offs across all cohorts

Next, we applied the cut-off values derived from the mpox survivor ROC curve to each cohort to evaluate the prevalence of individuals meeting the criteria for each antigen (Table 2). When using mpox-survivor–derived cut-offs, the known unexposed control group showed low-level background reactivity across several antigens (0–40%), consistent with nonspecific MSD signal rather than true OPXV exposure. In contrast, mpox survivors exhibited strong and broad seropositivity, with 76–90% reactivity across individual antigens. Composite metrics (all 5 VACV and all 5 MPXV) showed ∼90% seropositivity when at least one antigen in the five-marker VACV or MPXV panels exceeded its threshold, and 71–76% remained positive under the stricter rule requiring all five antigens to meet their respective cut-offs. Performance of the top three discriminative antigens (MPXV A35R, MPXV E8L, MPXV B6R; VACV A33R, VACV D8L, VACV B5R; Table 2 and Figure 3) was similarly high, with ∼85%-90% positivity when any one antigen was reactive and ∼85% when all three were required. All other exposed groups showed low reactivity. However, the MVA-BN vaccine group showed a strong response to VACV D8L, reaching 90.91% when measured on its own and performing significantly in the top VACV composites.

**Table 2:** Antigen-Specific Seroreactivity (%) Across Cohorts Using Mpox-Survivor Derived Cut-offs.

|  | Control<br>(n=22) | JJ EBOV +<br>MVA Boost<br>(n=22) | Jynneos<br>Vaccine<br>(n=21) | Mpox<br>Survivor<br>(n=21) | Smallpox<br>Survivor +<br>Vaccine<br>(n=22) | Smallpox<br>Vaccine<br>(n=25) |
| --- | --- | --- | --- | --- | --- | --- |
| VACV A27L | 27.27 | 9.09 | 27.27 | 76.19 | 40.91 | 20.00 |
| MPXV A29L | 36.36 | 36.36 | 40.91 | 85.71 | 63.64 | 40.00 |
| VACV A33R | 4.55 | 27.27 | 45.45 | 85.71 | 50.00 | 44.00 |
| VACV B5R | 0.00 | 9.09 | 18.18 | 85.71 | 36.36 | 24.00 |
| VACV D8L | 18.18 | 22.73 | 90.91 | 90.48 | 63.64 | 28.00 |
| VACV L1R | 9.09 | 18.18 | 9.09 | 80.95 | 27.27 | 8.00 |
| MPXV M1R | 9.09 | 18.18 | 18.18 | 76.19 | 40.91 | 16.00 |
| MPXV E8L | 4.55 | 4.55 | 54.55 | 85.71 | 22.73 | 4.00 |
| MPXV B6R | 0.00 | 4.55 | 18.18 | 85.71 | 40.91 | 28.00 |
| MPXV A35R | 4.55 | 27.27 | 13.64 | 85.71 | 50.00 | 44.00 |
| Composite 5<br>VACV | 40.91 | 45.45 | 90.91 | 90.48 | 77.27 | 52.00 |
| Composite 5<br>MPXV | 40.91 | 54.55 | 68.18 | 85.71 | 72.73 | 52.00 |
| Composite 5<br>VACV All | 0.00 | 0.00 | 0.00 | 71.43 | 18.18 | 8.00 |
| Composite 5<br>MPXV All | 0.00 | 4.55 | 0.00 | 76.19 | 18.18 | 4.00 |
| Top 3<br>VACV | 18.18 | 36.36 | 90.91 | 90.48 | 63.64 | 48.00 |
| Top 3<br>MPXV | 9.09 | 27.27 | 59.09 | 85.71 | 50.00 | 44.00 |
| Top 3<br>VACV All | 0.00 | 9.09 | 9.09 | 85.71 | 36.36 | 20.00 |
| Top 3<br>MPXV All | 0.00 | 4.55 | 0.00 | 85.71 | 22.73 | 4.00 |
| Top 6<br>VACV & MPXV | 22.73 | 36.36 | 90.91 | 90.48 | 63.64 | 48.00 |
| Top 6<br>VACV & MPXV All | 0.00 | 4.55 | 0.00 | 85.71 | 18.18 | 0.00 |

The seroreactivity status of all participants classified within the mpox survivor cohort is provided in Table S3. Diagnostic cutoff values for the three selected MPXV antigens (A35R, E8L, and B6R) were obtained from ROC curve analysis, which optimized sensitivity and specificity for distinguishing true positives from uninfected controls. These thresholds were subsequently applied to each participant’s quantified antibody concentration (AU/mL) to determine seropositive or seronegative status. Among the five MPXV antigens included in the MSD Orthopoxvirus panel, A35R, E8L, and B6R demonstrated the strongest discriminative performance, exhibiting the highest combined sensitivity and specificity in the mpox survivor cohort (Table 2).

All three of our generated ROC curves were used to compare potential cut-offs for each group (Table S1). To identify the most appropriate and significant cut-offs for our cohorts in the DRC, we first used the mpox survivor group (in green) as the true positive to evaluate specificity and sensitivity against the control group (Figure 2). The same ROC analysis was then conducted using the smallpox survivor and vaccinated group (in purple) as the true positive against the true negative controls (Figure S1).

**Figure 2:**
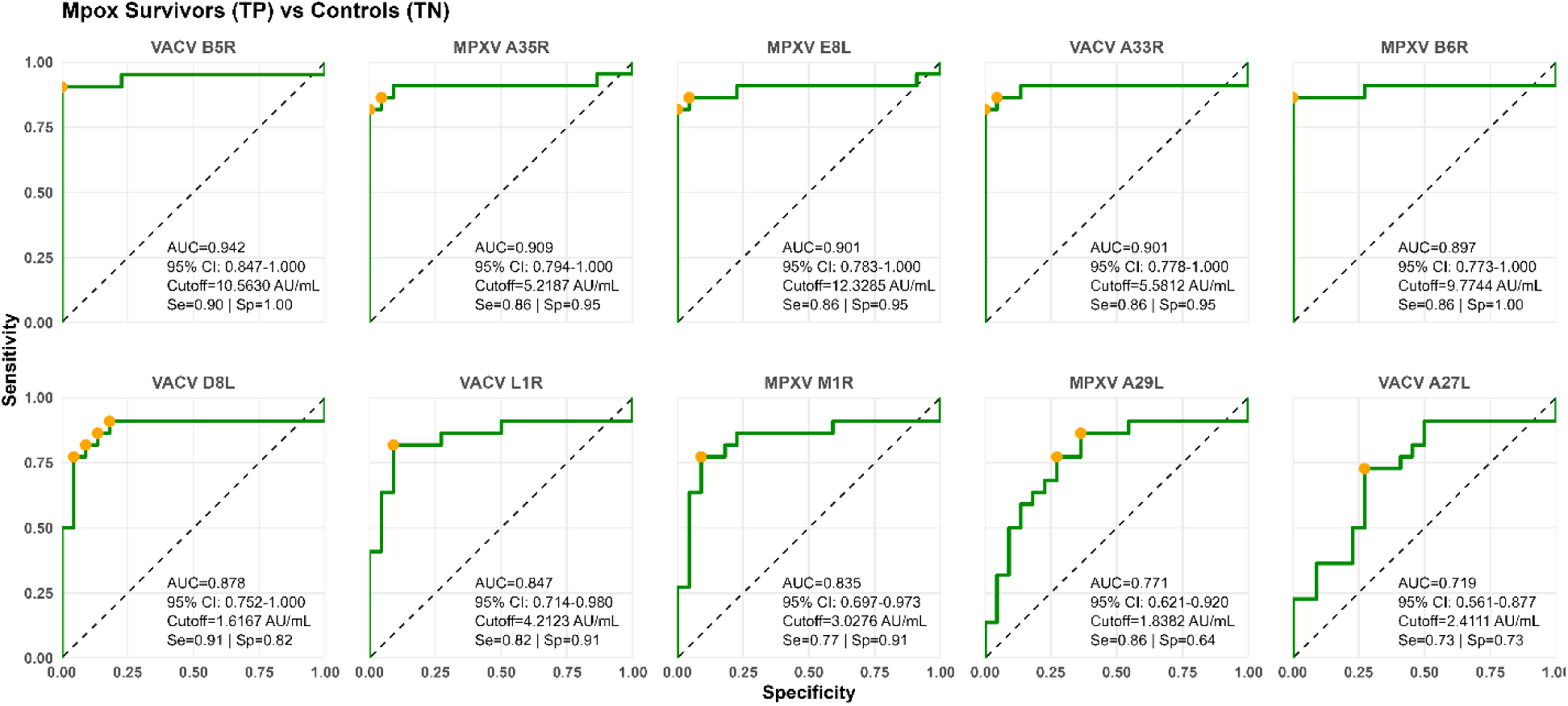
Receiver operating characteristic (ROC) curves comparing antibody responses in the mpox survivor cohort.

**Figure 3:**
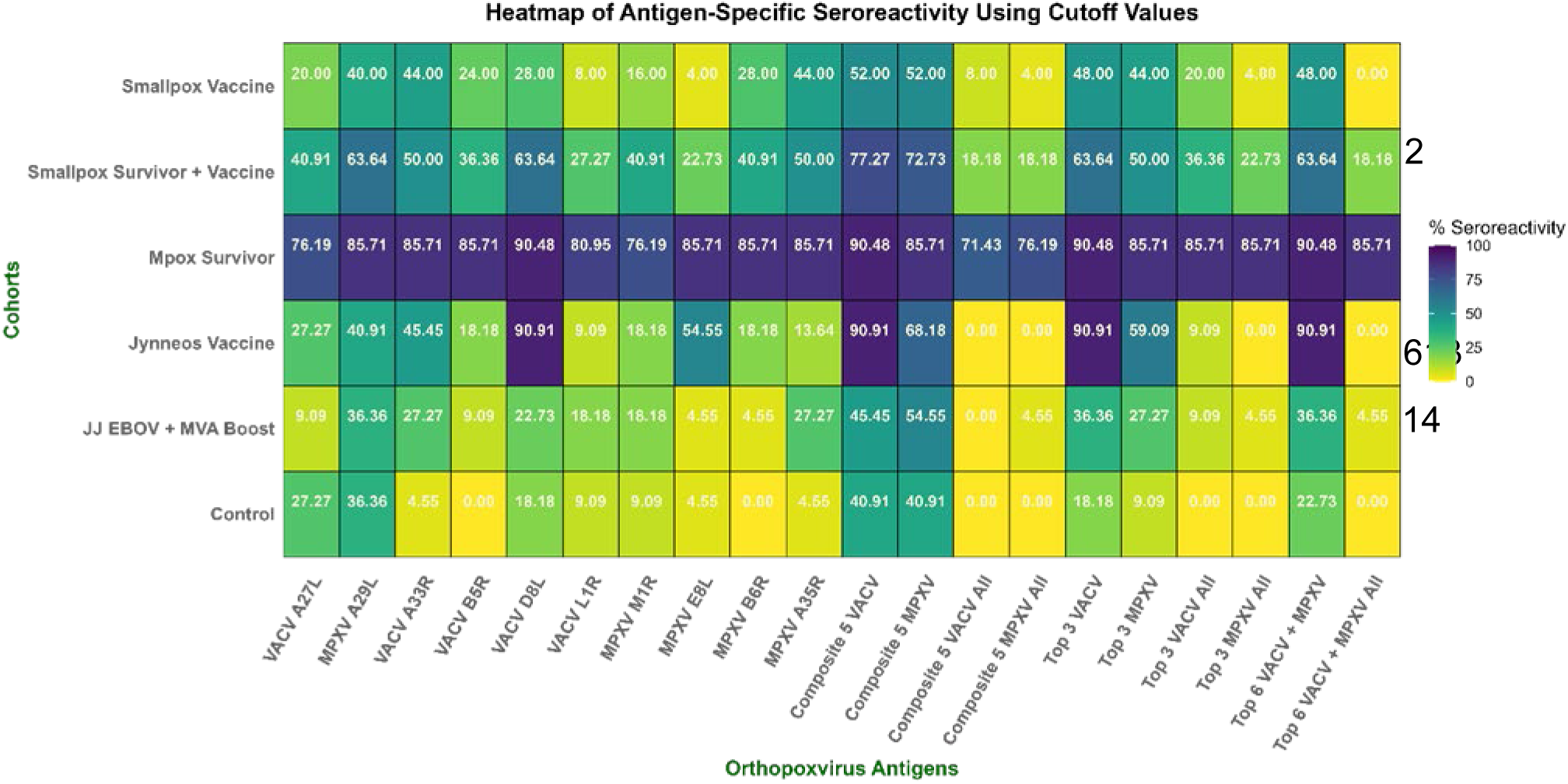
Heatmap of antigen-specific seroreactivity (%) across cohorts using mpox survivor derived cut-offs.

This cohort is composed of individuals with documented smallpox and who were subsequently immunized with Dryvax. The results shown reflect decades of persistent seroreactivity (Figure S1). The highest discriminatory power was found for VACV A33R (AUC = 0.64; CI = 0.51-0.77; Se = 0.48; Sp = 0.95; cutoff = 5.62 AU/mL), followed by VACV D8L (AUC = 0.62; CI = 0.48-0.76; Se = 0.53; Sp = 0.82; cutoff = 1.44 AU/mL), both exhibiting measurable antibody persistence long after eradication of the disease. L1R showed moderate diagnostic accuracy (AUC ≈ 0.56), while A27L displayed minimal separation from unexposed controls (AUC = 0.45), likely indicating a decline in antibody titers over time. Together, these findings reveal that A33R and D8L remain reliable indicators of long-term orthopoxvirus immunity, capturing the enduring humoral memory associated with historical smallpox exposure and vaccination.

Finally, we evaluated an ROC analysis for the combined groups: mpox survivors with smallpox survivors and vaccinated cohorts (in blue) to assess specificity and sensitivity across orthopoxviruses (Figure S2). This integrated analysis revealed a strong and consistent cross-reactive serological signature across both VACV and MPXV antigens. VACV A33R (AUC = 0.82; CI = 0.72-0.93; Se = 0.67; Sp = 0.95; cutoff = 5.58 AU/mL) and VACV D8L (AUC = 0.84; CI = 0.73-0.94; Se = 0.78; Sp = 0.86; cutoff = 2.80 AU/mL) demonstrated the highest overall diagnostic accuracy, highlighting their stability as long-term serological markers. Among MPXV antigens, A35R (AUC = 0.83; CI = 0.73-0.93; Se = 0.77; Sp = 0.91) and E8L (AUC = 0.76; CI = 0.65-0.88; Se = 0.67; Sp = 0.91) maintained good discriminatory performance, underscoring their value for detecting recent or cross-species orthopoxvirus exposure. Moderate separation was observed for B5R and L1R (AUCs > 0.80), while M1R and A29L displayed lower but reproducible distinction, suggesting antigen-specific differences in antibody longevity and epitope conservation. These results identified A33R, D8L, A35R, and E8L as the most robust composite markers of orthopoxvirus seroreactivity, bridging historical smallpox immunity with contemporary MPXV exposure and supporting their potential use as reference antigens.

### Performance of antigen panels in detecting MPXV and VACV exposures

We next assessed ROC performance for each antigen panel when applying the previously established seroreactivity cut-offs (Table 2). Across the mpox survivor cohort, all composite panels demonstrated high diagnostic accuracy, with AUC values ranging from 0.87 to 0.90. Panels built from the highest-performing antigens (MPXV A35R, MPXV E8L, MPXV B6R; VACV A33R, VACV D8L, VACV B5R) consistently showed strong sensitivity and specificity. Whether applying the “one-antigen-seroreactive” rule or requiring all antigens in the panel to exceed the cut-off, these markers reliably captured the most robust mpox-specific antibody responses. Importantly, looking at MPXV-specific antigen panels, Top 3 MPXV and Top 3 MPXV all, both present a very high AUC Se/Sp; (AUC= 0,88, Se= 0,81, Sp= 1.00) and (AUC=0.90, Se=0.86, Sp=96), which makes them two strong candidates to retain when establishing the appropriate cut-offs for specificity.

### Ortholog Ratios

The ratios between MPXV and VACV orthologs calculated from raw ECL signals (Figure S3) measured in the MSD assays are summarized in Table S2. In the mpox survivor group, the top antigens MPXV A35R (5.79 × 10^-5^), MPXV B6R (3.68 × 10^-6^), and MPXV E8L (0.0262) display significantly higher ratios compared with their VACV orthologs, highlighting their strong discriminatory potential despite high genetic similarity.

## DISCUSSION

In this study, we established individual antigen-specific cut-off values derived from our mpox survivor cohort. All serum samples were analyzed using the MSD Orthopoxvirus Serological Panel [25]. Antigen-specific cut-offs were determined using ROC curve analysis to identify values that maximize both sensitivity and specificity. These cut-offs enable differentiation between individuals who can be considered mpox seroreactive to the antigens included on the panel [26]. This classification framework provides a clear and consistent way to examine differences in antigen-specific antibody reactivity across cohorts with distinct OPXV exposure or vaccination profiles. These data formed the analytical basis for defining baseline seronegative and seropositive thresholds on the MSD platform [25, 26].

All cohorts included in this analysis were derived from a previously established sera panel collected in the DRC. The mpox survivor cohort was used to establish the antigen-specific cut-offs based on confirmed natural infection and documented histories indicating no prior vaccination or other orthopoxvirus exposure. The panel also included individuals representing multiple orthopoxvirus exposure groups, allowing antibody responses to be evaluated across diverse vaccination and exposure histories.

Our results indicate that the mpox antigens with the highest sensitivity and specificity were MPXV A35R, MPXV B6R, and MPXV E8L. Using the newly established cut-offs, the heatmap in Figure 4 shows a clear separation between mpox survivors and all other individuals in the study. Although a small percentage of signals remain in some grids, these are likely to reflect background noise or minimal cross-reactivity. Despite the generally high antigen similarity among OPXV, these results demonstrate that the established cut-offs can still effectively differentiate mpox-specific responses from other OPXV exposures.

On a molecular level, both A35R and B6R are surface proteins expressed on the EEV, which may contribute to higher antibody responses in mpox-infected individuals due to increased immune accessibility. In contrast, E8L is involved in viral entry, which may also explain its immunogenicity during natural infection [24, 27]. These studies consistently report that individuals with natural mpox infection exhibit higher antibody responses to E8L compared with negative controls, with some studies also reporting B6R as another antigen with high seroreactivity [20, 23]. Recent literature reports that the top three antigens with significant antibody responses to natural mpox infection are A35R, B6R, and E8L [5, 24]. In our analysis, we observed that the combination of the top three performing antigens (A35R, B6R, and E8L) also yields a higher percentage of seroreactivity. Adding a refined discriminatory rule to our binary composite, such as “if one of the three antigens meets the required cut-off value, consider it seroreactive,” helps identify potential mpox survivors or mpox-infected individuals.

With this rule requiring that all three antigens must meet the defined cut-off value, a more stringent criterion is applied, allowing for more specific identification of mpox-positive samples while reducing the likelihood that samples from individuals with other vaccination histories are classified as seroreactive. This stricter discriminatory rule may therefore be preferable for future assay development, as it enables clearer differentiation between mpox survivors and individuals with other OPXV vaccination backgrounds. As shown in Table 3, although the specificity is slightly lower (0.96), this approach yields higher overall diagnostic performance with an AUC of 0.90.

**Table 3:** ROC Analysis of Composite Antigen Panels for Mpox Survivors Cohort.

| Panel | AUC | Cut off (AU/mL) | Sensitivity | Specificity |
| --- | --- | --- | --- | --- |
| Composite 5 VACV | 0.87 | 51.71 | 0.81 | 0.96 |
| Composite 5 MPXV | 0.87 | 115.05 | 0.81 | 1.00 |
| Composite 5 VACV All | 0.89 | 34.62 | 0.81 | 1.00 |
| Composite 5 MPXV All | 0.89 | 7.53 | 0.86 | 0.96 |
| Top 3 VACV | 0.88 | 10.56 | 0.86 | 0.91 |
| Top 3 MPXV | 0.88 | 115.05 | 0.81 | 1.00 |
| Top 3 VACV All | 0.89 | 56.02 | 0.81 | 1.00 |
| Top 3 MPXV All | 0.90 | 10.24 | 0.86 | 0.96 |
| Top 6 VACV + MPXV | 0.88 | 161.90 | 0.81 | 1.00 |
| Top 6 VACV + MPXV All | 0.89 | 8.12 | 0.86 | 0.96 |

While VACV panel in mpox survivors exhibits slightly higher overall accuracy, the Top 3 MPXV panel demonstrates higher specificity, which is particularly valuable for identifying serum samples to be screened in future studies. These discriminatory rules help reduce false positives and will serve as the common parameters for further studies conducted in the DRC.

In other words, these cut-offs may enable the assay to selectively detect mpox-specific antibody responses while minimizing false-positive signals from VACV exposure or unexposed individuals (Table 4).

**Table 4:** Top 3 high-performing antigens and their established discriminatory cut-off values for MPXV serological analysis.

| Top 3<br>Composites | Antigen | Cut off (AU/mL) |
| --- | --- | --- |
|  | MPXV A35R | 5.22 |
|  | MPXV E8L | 12.33 |
|  | MPXV B6R | 9.77 |

While the MSD assay remains a relatively new tool for serological diagnostics and has yet to establish a gold standard cut-off, it represents a rapid and efficient approach for assessing immune responses, particularly in asymptomatic or post-infected individuals [20]. Several studies have evaluated its use in vaccinated populations to establish antigen-specific cut-offs and appropriate seroreactivity thresholds. Previous studies using multiplex orthopoxvirus serology have demonstrated that combining responses to multiple antigens can substantially improve the ability to distinguish MPXV infection from vaccination-induced immunity [5, 24]. In these analyses, highly reactive antigens such as E8, A35/A33, and B6/B5 contributed strongly to discrimination between serological groups, supporting the use of multi-antigen approaches rather than relying on a single marker [5, 20]. Although machine-learning models further improved classification accuracy, the results also highlight that carefully selected antigen combinations and defined decision rules can effectively capture key patterns in orthopoxvirus antibody responses [5, 24]. Consistent with this, multiplex orthopoxvirus serological assays show that natural mpox infection is associated with broader antibody reactivity across multiple viral antigens, whereas vaccinia vaccination typically induces more restricted recognition patterns, including antibodies targeting conserved orthopoxvirus surface proteins such as A35R and H3L, which were included in multiplex antigen panels to characterize infection-associated antibody profiles [28].

However, antigen-specific cut-off concentrations vary across studies, likely reflecting differences in exposure history, infection severity, occupational risk, and living conditions. Our dataset is unique in that it exclusively includes individuals residing in an active mpox-endemic region, providing valuable insight into naturally acquired immune responses. It is also important to note that strong antibody signals may be observed in smallpox survivors or vaccinated individuals. Notably, VACV antibodies have been reported to persist for up to 80 years [21]. Although our study did not focus on smallpox-specific infection, we generated ROC curves to explore cut-offs in smallpox survivor groups and in a combined mpox and smallpox survivor group (Figures S1, S2, and Table S1), which yielded slightly less accurate ROC curves.

### Limitations

Earlier investigations indicate that antigens such as A29L/A27L and M1R/L1R may exhibit stronger reactivity in vaccinated populations [20]. In this sample set, further analyses should include evaluating cut-offs for vaccinated individuals or stratifying groups by demographics such as age, sex, and infection history to assess significant variations in cut-offs. With ongoing surveillance across the DRC and continuous case reporting, it is important to monitor changes in incidence and the factors driving mpox expansion. Certainly, the small sample size cannot capture the full picture of mpox burden across Central Africa; however, these cut-offs provide a valuable starting point in these community settings.

### Future Directions

Future work should include neutralization assays to identify antigens that could still induce functional immune responses. Although this subset of individuals was specifically selected to evaluate proper cut-off values for discriminating mpox-specific exposures from other groups, performing MSD assays with serially diluted samples and expanding sample size across multiple settings would help establish more robust cut-off values, particularly in high-risk populations. Focusing on A35R and B6R in neutralization assays is crucial, as these surface proteins play key roles in mpox infection and are known for their neutralizing abilities [27, 29]

## CONCLUSION

Although mpox is no longer classified as a PHEIC, it remains an important high-risk pathogen. With the evolving dynamics of mpox infection in endemic communities and the adoption of new serologic screening tools to enhance surveillance, establishing accurate analytical thresholds remains critical for reliable detection and monitoring of the pathogen.

## Data Availability

All data produced in the present study are available upon reasonable request to the authors.

## ACKNOWLEDGEMENTS

This work was supported by the Gates Foundation under INV-085623, and through funding from the Canadian Institutes of Health Research and International Development Research Centre for the International Mpox Research Consortium (IMReC) (grant no. 202209MRR-489062-MPX-CDAA-168421).

## Supplementary Tables and Figures

**Figure S1:**
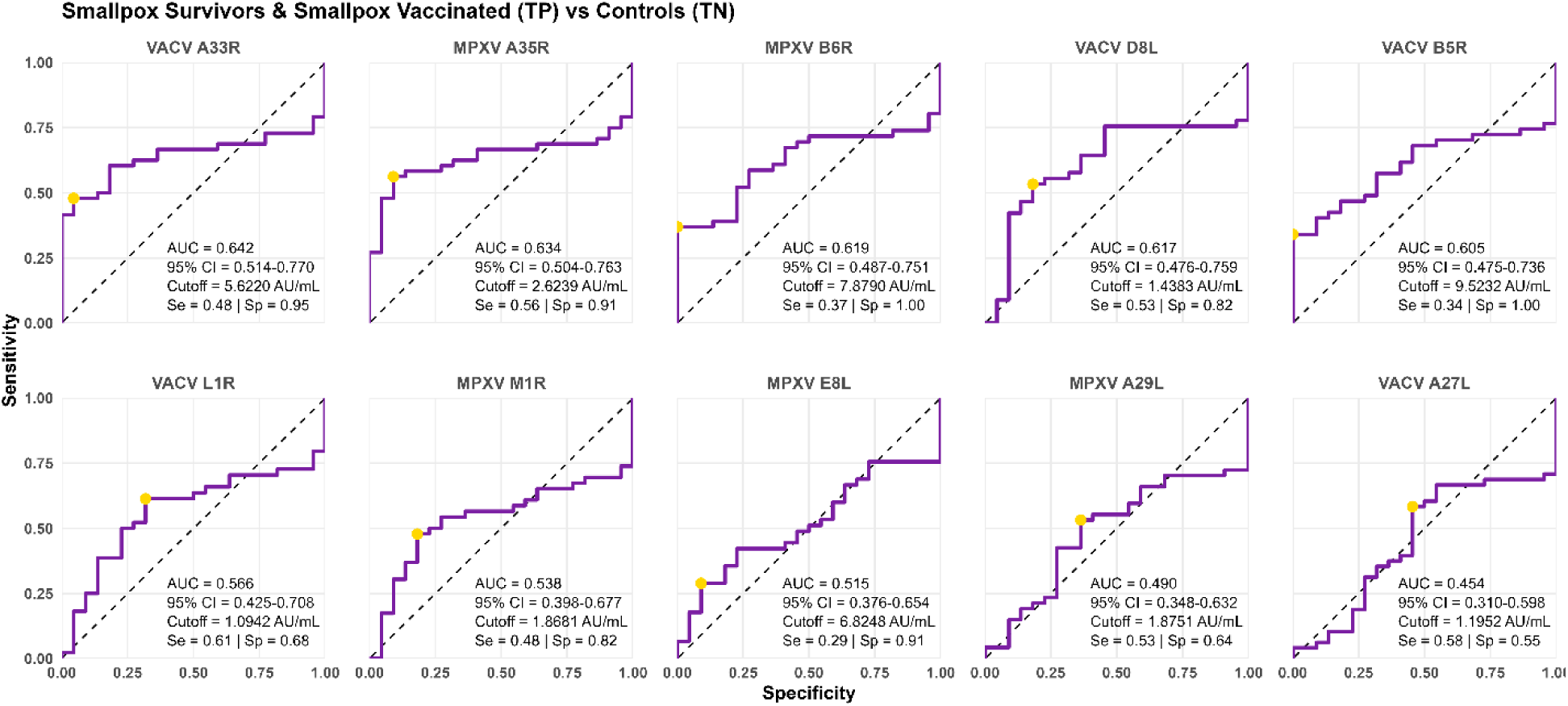
Receiver operating characteristic (ROC) curves comparing antibody responses in Smallpox & Vaccinated Survivors (true positives) versus controls (true negatives)

**Figure S2:**
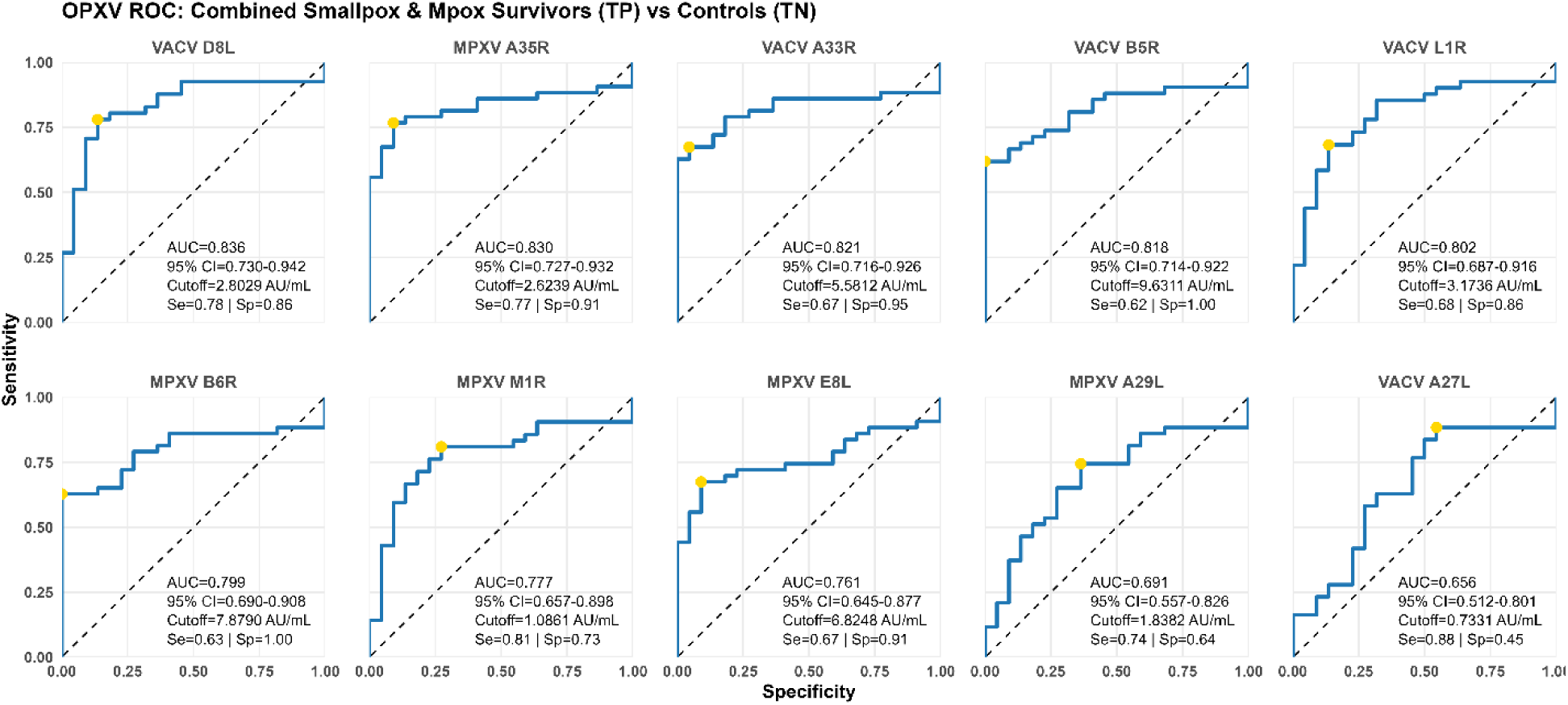
Receiver operating characteristic (ROC) curves combining antibody responses in smallpox + vaccinated survivors and mpox survivors (true positives) versus uninfected controls (true negatives).

**Figure S3:**
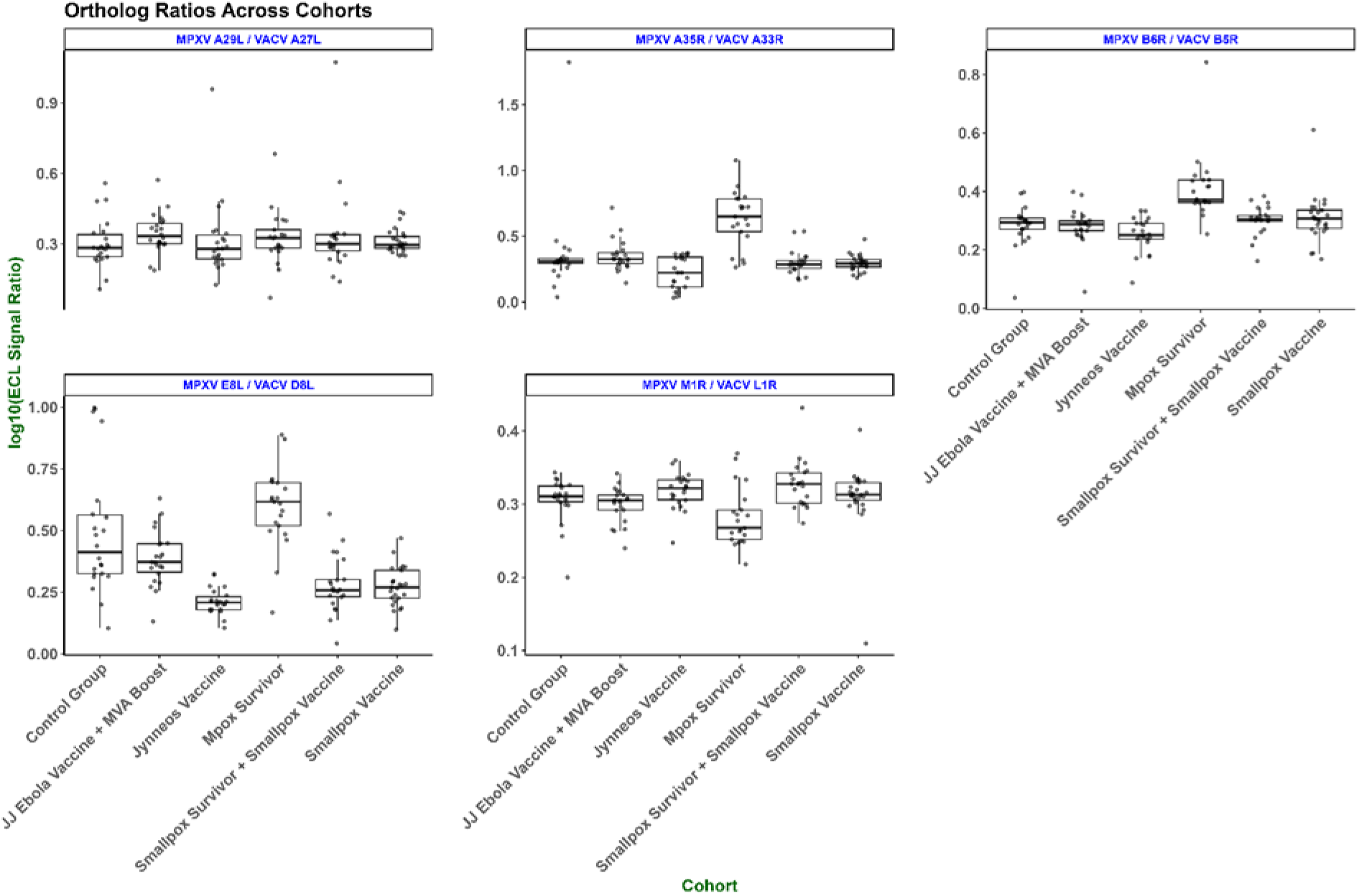
MPXV/VACV ortholog signal ratios (log₁₀ ECL) across participants.

**Table S1:** Summary of ROC analysis across mpox survivors, smallpox survivors, vaccinated Cohorts, and combinations thereof.

| Cohort | Antigen | AUC | Cut-off (AU/mL) | Sensitivity | Specificity |
| --- | --- | --- | --- | --- | --- |
| **Mpox Survivors** | MPXV A35R | 0.91 | 5.22 | 0.86 | 0.96 |
|  | VACV A33R | 0.90 | 5.58 | 0.86 | 0.96 |
|  | MPXV B6R | 0.90 | 9.77 | 0.86 | 1.00 |
|  | VACV B5R | 0.94 | 10.56 | 0.91 | 1.00 |
|  | VACV D8L | 0.88 | 1.62 | 0.91 | 0.82 |
|  | MPXV E8L | 0.90 | 12.33 | 0.86 | 0.96 |
|  | MPXV M1R | 0.84 | 3.03 | 0.77 | 0.91 |
|  | VACV L1R | 0.85 | 4.21 | 0.82 | 0.91 |
|  | MPXV A29L | 0.77 | 1.84 | 0.86 | 0.64 |
|  | VACV A27L | 0.72 | 2.41 | 0.73 | 0.73 |
| **Smallpox Survivors & Vaccinated** | MPXV A35R | 0.63 | 2.62 | 0.56 | 0.91 |
|  | VACV A33R | 0.64 | 5.62 | 0.48 | 0.96 |
|  | MPXV B6R | 0.62 | 7.88 | 0.37 | 1.00 |
|  | VACV B5R | 0.61 | 9.52 | 0.34 | 1.00 |
|  | MPXV E8L | 0.52 | 6.83 | 0.29 | 0.91 |
|  | VACV D8L | 0.62 | 1.44 | 0.53 | 0.82 |
|  | MPXV M1R | 0.54 | 1.87 | 0.48 | 0.82 |
|  | VACV L1R | 0.57 | 1.09 | 0.61 | 0.68 |
|  | MPXV A29L | 0.49 | 1.88 | 0.53 | 0.64 |
|  | VACV A27L | 0.45 | 1.20 | 0.58 | 0.55 |
| **Combined OPXV Exposures** | MPXV A35R | 0.83 | 2.62 | 0.77 | 0.91 |
|  | VACV A33R | 0.82 | 5.58 | 0.67 | 0.96 |
|  | MPXV B6R | 0.80 | 7.88 | 0.63 | 1.00 |
|  | VACV B5R | 0.82 | 9.63 | 0.62 | 1.00 |
|  | MPXV E8L | 0.76 | 6.83 | 0.67 | 0.91 |
|  | VACV D8L | 0.84 | 2.80 | 0.78 | 0.86 |
|  | MPXV M1R | 0.78 | 1.09 | 0.81 | 0.73 |
|  | VACV L1R | 0.80 | 3.17 | 0.68 | 0.86 |
|  | MPXV A29L | 0.69 | 1.84 | 0.74 | 0.64 |
|  | VACV A27L | 0.66 | 0.73 | 0.88 | 0.46 |

**Table S2:**
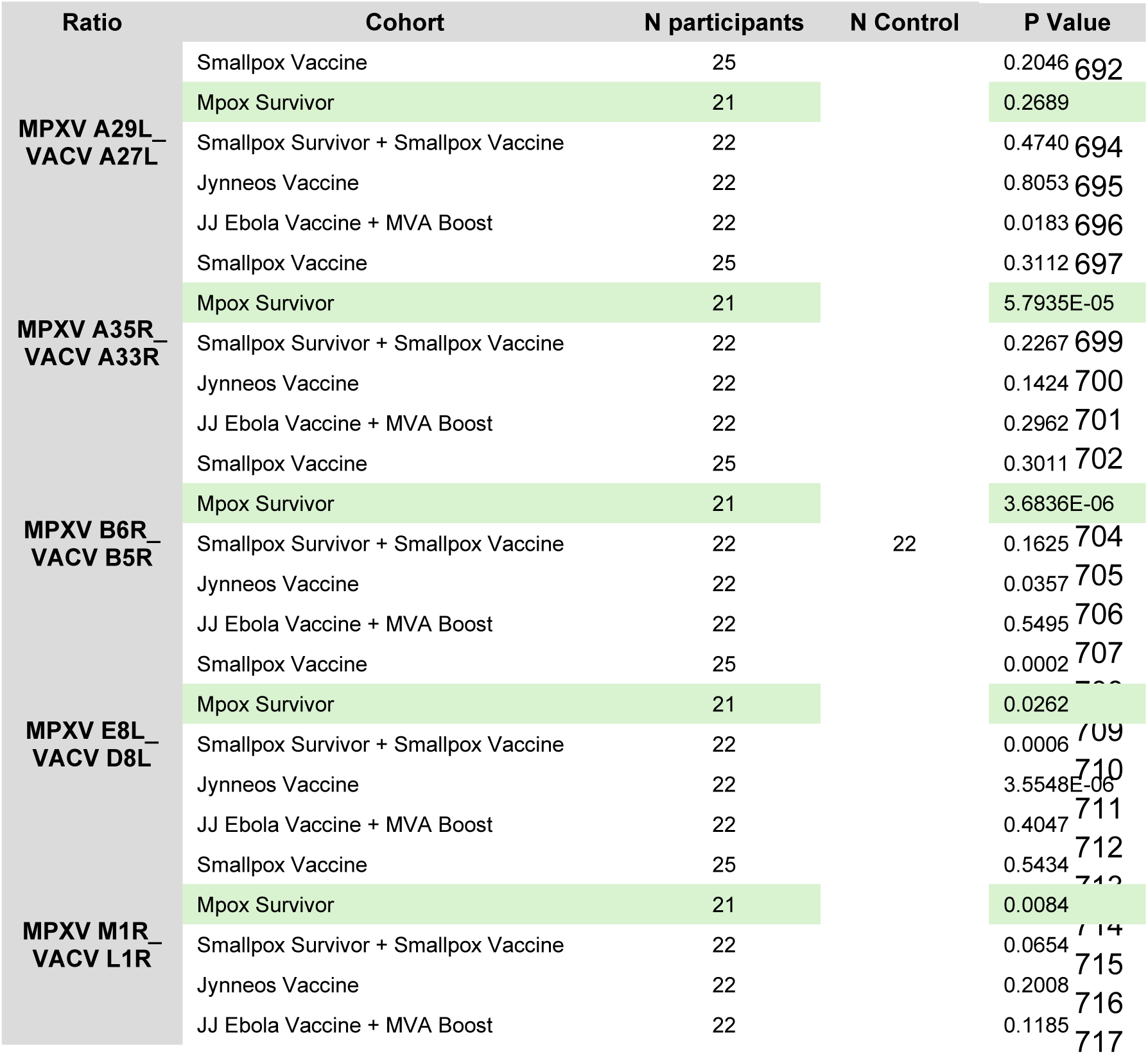
Wilcoxon test results for MPXV/VACV ortholog ratios across cohorts. P-values were calculated using the Wilcoxon test to assess differences in antigen-binding efficiency between the ortholog pairs.

**Table S3:**
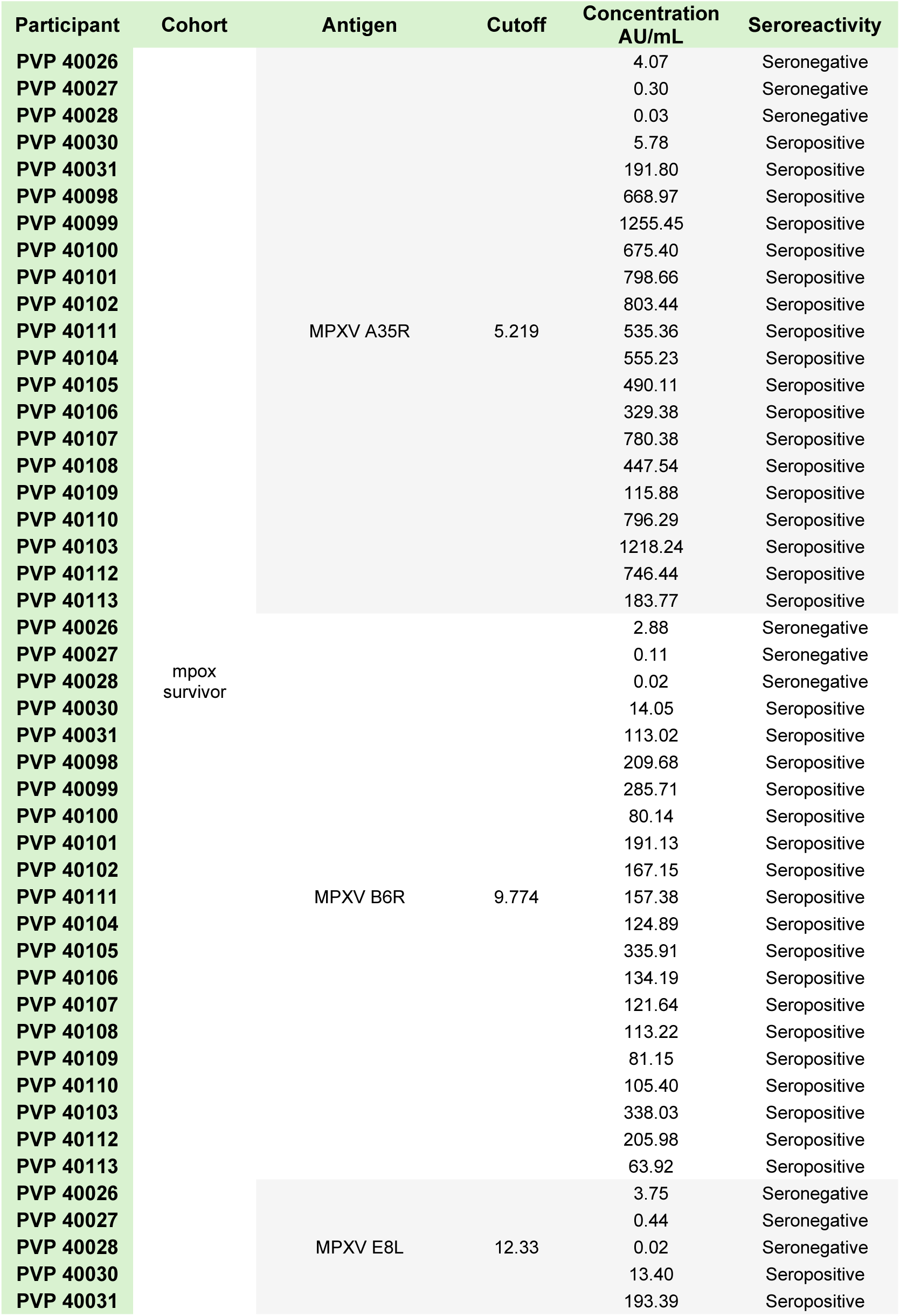
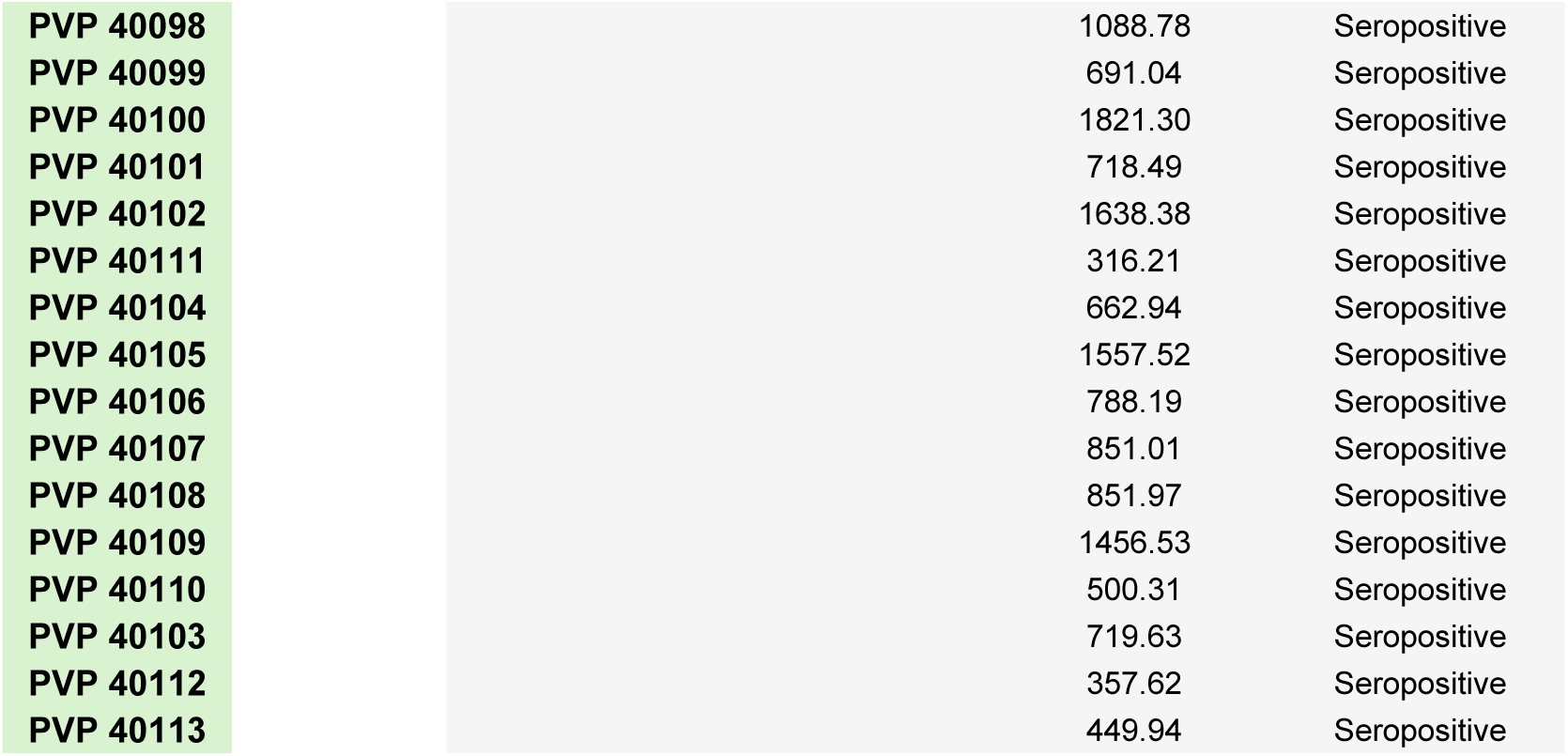
Seroreactivity classification across three MPXV antigens in mpox survivors using antigen-specific diagnostic thresholds.

## REFERENCES

1. Huang, Y., L. Mu, and W. Wang, Monkeypox: epidemiology, pathogenesis, treatment and prevention. Signal Transduction and Targeted Therapy, 2022. 7(1): p. 373.

2. Hutson, C.L., et al., Comparison of Monkeypox Virus Clade Kinetics and Pathology within the Prairie Dog Animal Model Using a Serial Sacrifice Study Design. Biomed Res Int, 2015. 2015: p. 965710.

3. Luna, N., et al., Phylogenomic analysis of the monkeypox virus (MPXV) 2022 outbreak: Emergence of a novel viral lineage? Travel Medicine and Infectious Disease, 2022. 49: p. 102402.

4. Wawina-Bokalanga, T., et al., *Co-circulation of monkeypox virus subclades Ia and Ib in Kinshasa Province, Democratic Republic of the Congo, July to* August *2024*. Eurosurveillance, 2024. 29(38): p. 2400592.

5. Surtees, R., et al., Machine learning-supported framework for the classification of mpox infection and MVA immunization from multiplexed serology data. Nature Communications, 2025. 16(1): p. 10943.

6. Kinganda-Lusamaki, E., et al., *Clade I mpox virus genomic diversity in the Democratic Republic of the Congo*, *2018-2024: Predominance of zoonotic transmission*. Cell, 2025. 188(1): p. 4-14.e6.

7. Organization, W.H. WHO Director-General declares mpox outbreak a public health emergency of international concern. 2026 [cited 2025 November 24th]; Available from: https://www.who.int/news/item/14-08-2024-who-director-general-declares-mpox-outbreak-a-public-health-emergency-of-international-concern.

8. Nizigiyimana, A., et al., *Epidemiological analysis of confirmed mpox cases, Burundi*, *3 July to 9 September 2024*. Eurosurveillance, 2024. 29(42): p. 2400647.

9. (WHO), W.H.O. Fifth meeting of the International Health Regulations (2005) Emergency Committee regarding the upsurge of mpox 2024. 2026 [cited 2026 February 20th]; Available from: https://www.who.int/news/item/30-10-2025-fifth-meeting-of-the-international-health-regulations-(2005)-emergency-committee-regarding-the-upsurge-of-mpox-2024.

10. Karagoz, A., et al., Monkeypox (mpox) virus: Classification, origin, transmission, genome organization, antiviral drugs, and molecular diagnosis. Journal of Infection and Public Health, 2023. 16(4): p. 531–541.

11. Tang, D., et al., Recombinant proteins A29L, M1R, A35R, and B6R vaccination protects mice from mpox virus challenge. Frontiers in Immunology, 2023. **Volume 14** - 2023.

12. Wahl, V., et al., Variola Virus and Clade I Mpox Virus Differentially Modulate Cellular Responses Longitudinally in Monocytes During Infection. J Infect Dis, 2024. 229(Supplement_2): p. S265–s274.

13. Grifoni, A., et al., Defining antigen targets to dissect vaccinia virus and monkeypox virus-specific T cell responses in humans. Cell Host & Microbe, 2022. 30(12): p. 1662–1670.e4.

14. Tan, C., et al., Development of multi-epitope vaccines against the monkeypox virus based on envelope proteins using immunoinformatics approaches. Frontiers in Immunology, 2023. **Volume 14** - 2023.

15. Greenberg, R.N. and J.S. Kennedy, ACAM2000: a newly licensed cell culture-based live vaccinia smallpox vaccine. Expert Opin Investig Drugs, 2008. 17(4): p. 555–64.

16. Mohamed Abdoul-Latif, F., et al., Mpox Resurgence: A Multifaceted Analysis for Global Preparedness. Viruses, 2024. 16(11).

17. (WHO), W.H.O. WHO adds LC16m8 mpox vaccine to Emergency Use Listing. 2024 [cited 2026 March 23rd]; Available from: https://www.who.int/news/item/19-11-2024-who-adds-lc16m8-mpox-vaccine-to-emergency-use-listing?utm_source=chatgpt.com.

18. Vakaniaki, E.H., et al., Sustained human outbreak of a new MPXV clade I lineage in eastern Democratic Republic of the Congo. Nature Medicine, 2024. 30(10): p. 2791–2795.

19. Kinganda-Lusamaki, E., et al., Use of Mpox Multiplex Serology in the Identification of Cases and Outbreak Investigations in the Democratic Republic of the Congo (DRC). Pathogens, 2023. 12(7): p. 916.

20. Reed, J.C., et al., Differentiating mpox infection and vaccination using a validated multiplex orthopoxvirus IgG serology assay. Journal of Clinical Microbiology, 2026. 64(2): p. e01548–25.

21. Cohn, H., et al., Mpox vaccine and infection-driven human immune signatures: an immunological analysis of an observational study. The Lancet Infectious Diseases, 2023. 23(11): p. 1302–1312.

22. Mazzotta, V., et al., Kinetics of the humoral and cellular immune response up to 1 year from mpox virus infection. Clinical Microbiology and Infection, 2025. 31(8): p. 1356–1362.

23. Hunt, J.H., et al., Discordant performance of mpox serological assays. Journal of Virological Methods, 2024. 329: p. 115004.

24. Pettke, A., et al., Serological differentiation between naturally acquired mpox and MVA-BN-vaccine induced antibody responses using ratios of MPXV and VACV antigen pairs in the MSD immunoassay. Microbiology Spectrum, 2025. 13(9): p. e00182–25.

25. (MSD), M.D. V-PLEX Orthopoxvirus Panel 1 (IgG) Kit. 2026 [cited 2025 October 16th]; Available from: https://www.mesoscale.com/en/products/v-plex-orthopoxvirus-panel-1-igg-kit-k15688u/.

26. Nahm, F.S., Receiver operating characteristic curve: overview and practical use for clinicians. Korean J Anesthesiol, 2022. 75(1): p. 25–36.

27. Wang, Y., K. Yang, and H. Zhou, Immunogenic proteins and potential delivery platforms for mpox virus vaccine development: A rapid review. Int J Biol Macromol, 2023. 245: p. 125515.

28. Yates, J.L., et al., Development of a novel serological assay for the detection of mpox infection in vaccinated populations. J Med Virol, 2023. 95(10): p. e29134.

29. Liu, J., et al., Immunogenicity of monkeypox virus surface proteins and cross-reactive antibody responses in vaccinated and infected individuals: implications for vaccine and therapeutic development. Infectious Diseases of Poverty, 2025. 14(1): p. 12.

